# Risk of long COVID and associated symptoms after acute SARS-COV-2 infection in ethnic minorities: a Danish nationwide cohort study

**DOI:** 10.1101/2023.08.22.23294402

**Authors:** George F. Mkoma, Charles Agyemang, Thomas Benfield, Mikael Rostila, Agneta Cederström, Jørgen H. Petersen, Marie Norredam

## Abstract

**Background:** Ethnic minorities living in high-income countries have been disproportionately affected by COVID-19 in terms of infection rates and hospitalisations; however, less is known about long COVID in this population. Our aim was to examine the risk of long COVID and associated symptoms among ethnic minorities.

**Methods and Findings:** A Danish nationwide register-based cohort study of individuals diagnosed with COVID-19 aged ≥18 years (n=2 334 271) between January 2020 and August 2022. We calculated the risk of long COVID diagnosis and long COVID symptoms among ethnic minorities compared with native Danes using multivariable Cox proportional hazard regression and logistic regression, respectively.

Ethnic minorities from North Africa (adjusted hazard ratio [aHR] 1.41; 95% CI 1.12–1.79), Middle East (aHR 1.38; 95% CI 1.24–1.55), Eastern Europe (aHR 1.35; 95% CI 1.22–1.49), and Asia (aHR 1.23; 95% CI 1.09–1.40) had significantly greater risk of long COVID diagnosis than native Danes in both unadjusted and adjusted models. In the analysis by largest countries of origin, the greater risks of long COVID diagnosis were found in Iraqis (aHR 1.56; 95% CI 1.30– 1.88), Turks (aHR 1.42; 95% CI 1.24–1.63), and Somalis (aHR 1.42; 95% CI 1.07–1.91) after adjustment for confounders. Significant factor associated with an increased risk of long COVID diagnosis was COVID-19 hospitalisation. Furthermore, the odds of reporting cardiopulmonary symptoms (including dyspnoea, cough, and chest pain) and *any* long COVID symptoms were higher among North African, Middle Eastern, Eastern European, and Asian than among native Danes in both unadjusted and adjusted models.

**Conclusions:** Belonging to an ethnic minority group was significantly associated with an increased risk of long COVID indicating the need to better understand long COVID drivers and address care and treatment strategies in this population.

## Introduction

Globally, millions of people have now been infected with SARS-COV-2, the virus causing coronavirus disease 2019 (COVID-19) [1]. Despite increased risk of hospitalisation and death in the first weeks of SARS-COV-2 infection, many COVID-19 survivors experience a range of symptoms including fatigue, cardiopulmonary symptoms (dyspnoea, cough, and chest pain), and neurological symptoms (headache, depression, and memory loss) persisting beyond weeks or months after the acute phase of COVID-19 infection; the condition known as long COVID as per National Institute for Health and Care Excellence (NICE) guidelines [2–5]. Long COVID or post-acute sequelae of COVID-19 is an emerging epidemic that is anticipated to affect the quality of life of many COVID-19 survivors [6,7]. Hence, understanding the demographic profile of long COVID sufferers is of use for planning healthcare services.

Ethnic minorities living in high-income countries have been disproportionately affected by COVID-19 in terms of infection rates, hospitalisations, and severe morbidity [8]. However, studies on long COVID among ethnic minorities are few and their findings suggest that this population exhibits a greater risk of long COVID [9–14]. For example, compared with the majority White populations in the United States and the United Kingdom, individuals who belong to Black and Asian ethnicity were observed to have a higher chance of reporting long COVID symptoms after acute COVID-19 infection [9–13]. In the Netherlands, the risk of long COVID was found to be higher in patients with Surinamese, Moroccan, and Turkish origin than in those with Dutch origin [14]. Overall, the previous studies have several shortcomings, including studies were based on a single hospital setting or localised area, the studies did not compare symptoms distribution before and after COVID-19 diagnosis, and most of the studies were survey-based. In addition, comorbidities and socioeconomic factors such as income and education were not considered in some studies, albeit these factors may impact the likelihood of reporting long COVID symptoms. Notwithstanding the limitations from the previous research, evidence has emerged showing that older age, disease severity, use of intensive care, comorbidities, and not receiving COVID-19 vaccine are associated with increased risk of long COVID in the general population [2,3,15].

In the present study, nationwide register data from individuals diagnosed with COVID-19 in Denmark were used. First, we hypothesised that ethnic minorities (defined by their region and country origin) have a higher risk of long COVID diagnosis compared to native Danes taking into account comorbidities, socioeconomic factors, civil status, COVID-19 related hospitalisation, and vaccination status. Second, we examined whether the risk of fatigue, headache, cardiopulmonary, or *any* of these long COVID symptoms differed between ethnic minorities and native Danes within 6 months before COVID-19 diagnosis, 0 to 4 weeks, and >4 weeks to 6 months after COVID-19 diagnosis.

## Methods

### Setting

Denmark has a population of approximately 5.8 million people. Testing for SARS-COV-2 infection by PCR was launched in March 2020. During March–May 2020, testing for SARS-COV-2 by PCR was offered for individuals with mild to severe symptoms of respiratory tract infection [16]. Universal testing for SARS-COV-2 infection by PCR was nationally implemented from May 18, 2020. Additionally, vaccination against COVID-19 started on December 27, 2020 [16]. The Danish healthcare system is publicly financed by general taxes and access to test for SARS-COV-2 and vaccination against COVID-19 is free of charge for all residents [17].

### Data sources and study population

This nationwide register-based cohort study utilised data from the Danish COVID-19 surveillance database, the Danish National Patient Registry (DNPR), the Danish Vaccination Register, and Statistics Denmark. The study population included all individuals residing in Denmark who had first-time tested positive for SARS-COV-2 (COVID-19 diagnosis) aged 18 years or older from January 1, 2020 to August 31, 2022 [18]. The study population was linked with the DNPR, which is a nationwide hospital register containing information on all primary and secondary diagnoses among hospitalised patients [19]. The DNPR contributed data on individuals who had COVID-19 as the primary reason for hospitalisation identified in accordance with 10^th^ version of International Standard Classification of Diseases (ICD-10): ICD-10 codes B34.2, B34.2A, B97.2, or B97.2A. Furthermore, the DNPR provided information on comorbidities and symptoms related to hospital contacts before and after COVID-19 diagnosis. We retrieved data on first, second, and third dose of COVID-19 vaccine from the Danish Vaccination Register [20]. Statistics Denmark contributed individual-level data on country of origin, date of immigration, highest attained education, family income, civil status, and date of death [21–23]. Linkage between the registers was possible due to the availability of unique personal identification number assigned to all Danish residents [23].

### Region and country of origin

The study population was categorised based on individual and parental region and country of origin [24]. The following eight groups were constructed according to their region of origin, with these groups being the modified version of those used by the World Bank: (i) Denmark (ii) Northern Europe, (iii) Western Europe, (iv) Eastern Europe, (v) Asia, (vi) Middle East, (vii) North Africa, and (viii) Subsaharan Africa [25]. Participants from North America, South America, and Oceania were excluded in the study as their numbers were relatively small. Furthermore, we classified the study population based on the largest countries of origin among the population of ethnic minorities residing in Denmark. The largest countries of origin selected were Norway, Sweden, Afghanistan, Iraq, Iran, Somalia, Pakistan, and Turkey. Individuals originating outside Denmark and their descendants formed the ethnic minority population. Participants originating and/or born in Denmark (native Danes) constituted the reference group.

### Outcome

The study participants were followed up from the date of a positive test for SARS-COV-2 infection until a long COVID diagnosis, death, emigration, or study end (August 31, 2022), whichever came first. The primary outcome of interest was long COVID diagnosis defined as complications persisting beyond the acute COVID-19 infection that cannot be explained by alternative diagnosis [26]. Presence of a long COVID diagnosis was determined by both ICD-10 codes (B94.8 or B94.8A) and the actual date of diagnosis. In addition, we examined hospital contacts related to long COVID symptoms such as fatigue, headache, dyspnoea, chest pain, cough, and depression or anxiety as a secondary outcome. Symptoms were identified by ICD-10 codes in relation to the date of hospital contact (Supplementary Table S1). Due to small outcome events on a single symptom by ethnic group, some symptoms were assessed as a composite outcome. In the present study, the following groups of symptoms were considered: fatigue, headache, cardiopulmonary symptoms (including dyspnoea, cough, and chest pain), and *any* of these selected long COVID symptoms. The analysis was restricted to the population experiencing these groups of symptoms within 6 months before COVID-19 diagnosis, 0 to 4 weeks (acute phase of COVID-19 infection), and >4 weeks to 6 months after COVID-19 diagnosis.

### Covariates

Covariates included in the analysis were age, sex, comorbidities, civil status, highest attained education, family income, length of residency, COVID-19 hospitalisation (as a proxy for disease severity), and vaccination against COVID-19. Age was analysed as a continuous variable and subsequently categorised as 18–60 years and >60 years in further analyses. COVID-19 hospitalisation was assessed as yes or no. Presence of comorbidities was determined by Charlson Comorbidity Index (CCI) based on discharge diagnosis within five years prior to COVID-19 diagnosis (Supplementary Table S1). The CCI included 17 conditions with scores assigned according to their severity [27]. The CCI score was divided into three groups: 0 (indicating no comorbidity), 1–2, and ≥3. Civil status was classified as cohabiting, living alone, or other. Education was grouped as low, medium, or high in accordance with the International Standard Classification of Education [28]. Income was categorised as low, middle, or high tertiles according to the total income distribution among patients with COVID-19 in the specific calendar year. Length of residency was a time difference in years between date of arrival in Denmark and date of COVID-19 diagnosis.

### Statistical analyses

Categorical and continuous variables were summarised by frequencies and percentages and by medians and interquartile ranges, respectively. We used multivariable Cox proportional hazard regression models to investigate the association between region and country of origin and the risk of long COVID diagnosis. Age, sex, civil status, education, family income, and CCI were identified as confounders using directed acyclic graphs; hence, these covariates were adjusted in the Cox models (Supplementary Figure S1). We refrained from adjusting for length of residency, COVID-19 hospitalisation, and COVID-19 vaccination status as these covariates were deemed to belong in the causal pathway for the risk of long COVID/reporting long COVID symptoms. The proportional hazard assumption was assessed by Schoenfeld residuals. In addition, we performed subgroup analyses in which the risk of long COVID diagnosis was compared between ethnic minorities and native Danes by age groups (18–60 years vs. >60 years), by COVID-19 hospitalisation (no vs. yes), and by COVID-19 vaccination (yes vs. no). The native Danes aged 18–60 years, native Danes non-hospitalised, and native Danes vaccinated were the reference groups in the three subgroup analyses.

Furthermore, we assessed the association between region and country of origin and hospital contacts related to groups of symptoms by fitting multivariable logistic regression models using the same set of covariates like in the Cox models. We compared hospital contacts related to groups of symptoms within 6 months before vs. 6 months after COVID-19 diagnosis in each ethnic group. Subsequently, we analysed hospital contacts related to groups of symptoms comparing ethnic minorities and native Danes in three time periods: 6 months before COVID-19 diagnosis, 0 to 4 weeks, and >4 weeks to 6 months after COVID-19 diagnosis. All hazard ratios (HR) and odds ratios (OR) with their corresponding 95% confidence interval (CI) were presented as unadjusted and adjusted, with native Danes regarded as the reference population. All analyses were performed in R statistical software (version 4.2.2).

## Results

### Participants characteristics

Between January 2020 and August 2022, 2 334 271 individuals were first-time diagnosed with COVID-19, of whom 40 321 (1.7%) were hospitalised and 2 292 950 (98.3%) were non-hospitalised individuals (Figure 1). Of the diagnosed COVID-19 cases, 1 973 998 (84.6%) were native Danes and 360 273 (15.4%) were ethnic minorities. Overall, 6479 native Danes and 755 ethnic minorities died within 6 months after COVID-19 diagnosis. The results on sociodemographic characteristics of the study participants showed that compared with native Danes, ethnic minorities, particularly those from Eastern Europe, Asia, Middle East, North Africa, and Subsaharan Africa were younger at the time of COVID-19 diagnosis and were more likely to have low level of education and more likely to have low family income (Table 1). North African (4.6%), Middle Eastern (4.2%), Eastern European (2.7%), Asian (2.8%), and Subsaharan African (2.5%) were in general more likely than native Danes (1.5%) to be hospitalised for COVID-19. Additionally, ethnic minorities from North Africa (28.5%) and Middle East (26.1%) as well as those from Pakistan (28.5%), Turkey (27.8%), Iraq (27.5%), Iran (26.2%), and Afghanistan (24.7%) had a higher proportion of individuals with comorbidities (CCI score of 1–2) than native Danes (20.3%) (Supplementary Table S2).

**Figure 1.**
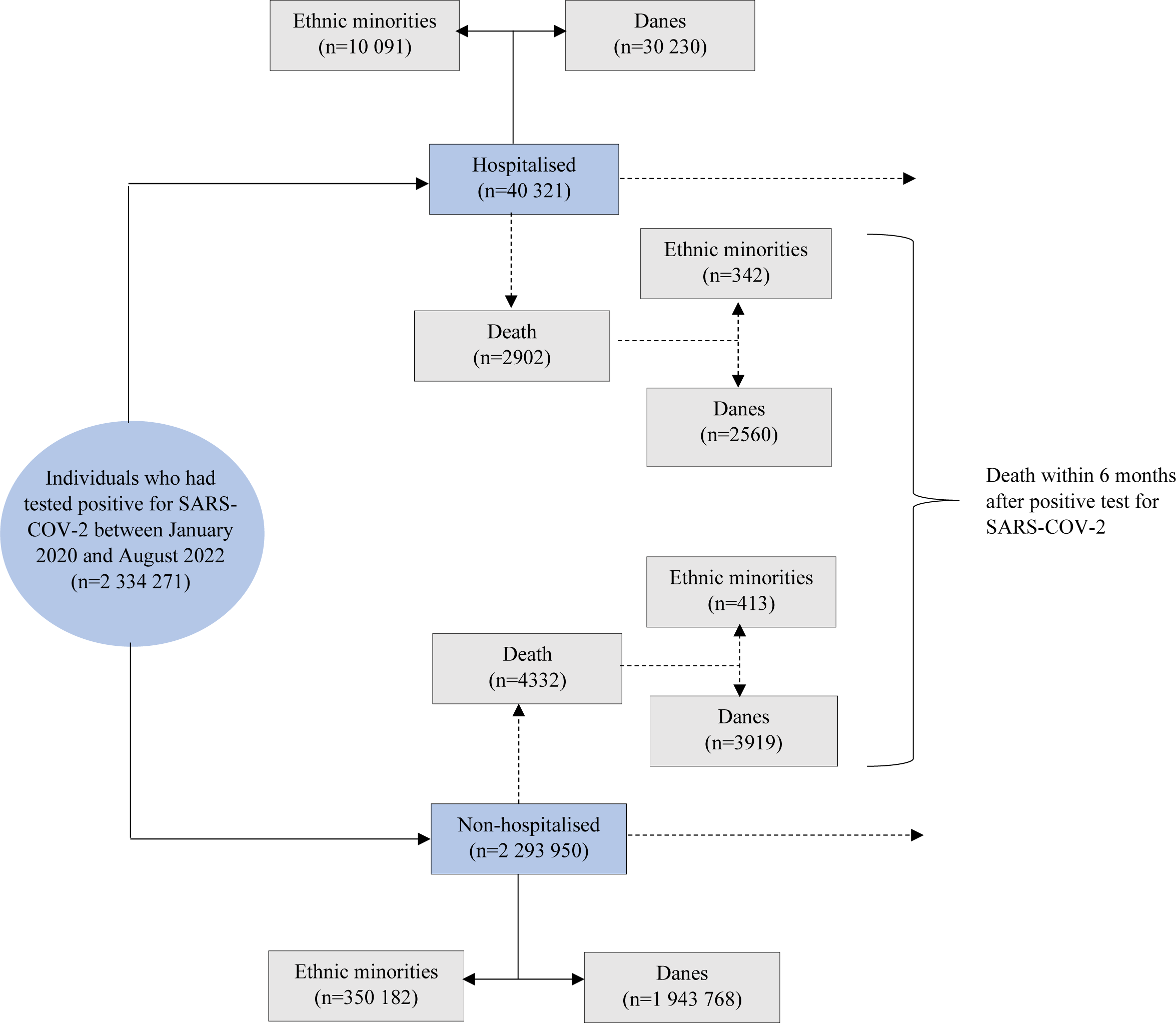
Flowchart of the study population.

**Table 1.**
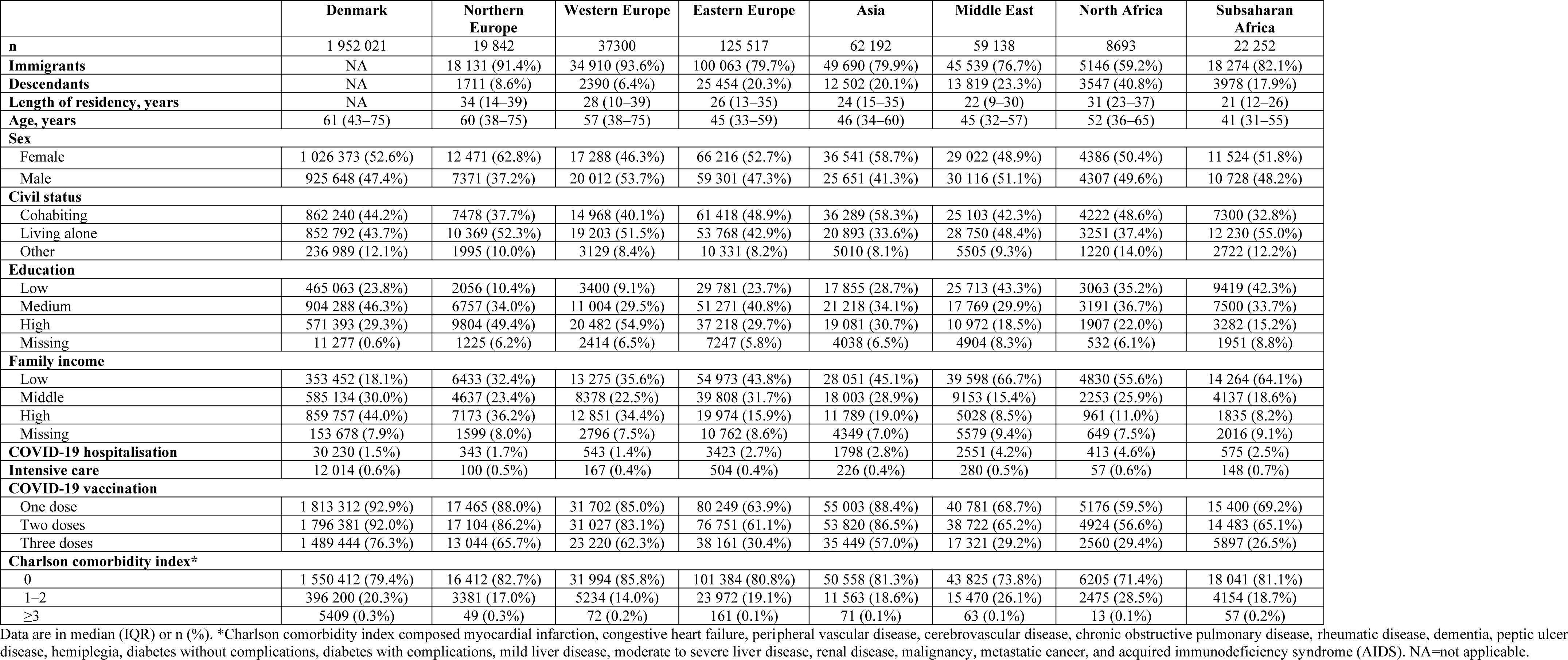
Individuals who had first-time tested positive for SARS-COV-2 between January 2020 and August 2022 by region of origin.

### Risk of long COVID diagnosis

We found that North African (HR 1.35; 95% CI 1.10–1.67), Middle Eastern (HR 1.31; 95% CI 1.18–1.44), Eastern European (HR 1.21; 95% CI 1.11–1.32), and Asian (HR 1.14; 95% CI 1.03–1.28) had a higher risk of long COVID diagnosis than native Danes in unadjusted model (Figure 2). After adjustment for age, sex, civil status, education, family income, and comorbidities, the risk of long COVID diagnosis remained significantly higher in North African (adjusted hazard ratio [aHR] 1.41; 95% CI 1.12–1.79), Middle Eastern (aHR 1.38; 95% CI 1.24–1.55), Eastern European (aHR 1.35; 95% CI 1.22–1.49), and Asian (aHR 1.23; 95% CI 1.09–1.40) than in native Danes. In the analysis by largest countries of origin, the results were most evident in individuals originating from Iraq (aHR 1.56; 95% CI 1.30–1.88), Turkey (aHR 1.42; 95% CI 1.24–1.63), and Somalia (aHR 1.42; 95% CI 1.07–1.91) (Figure 3).

**Figure 2.**
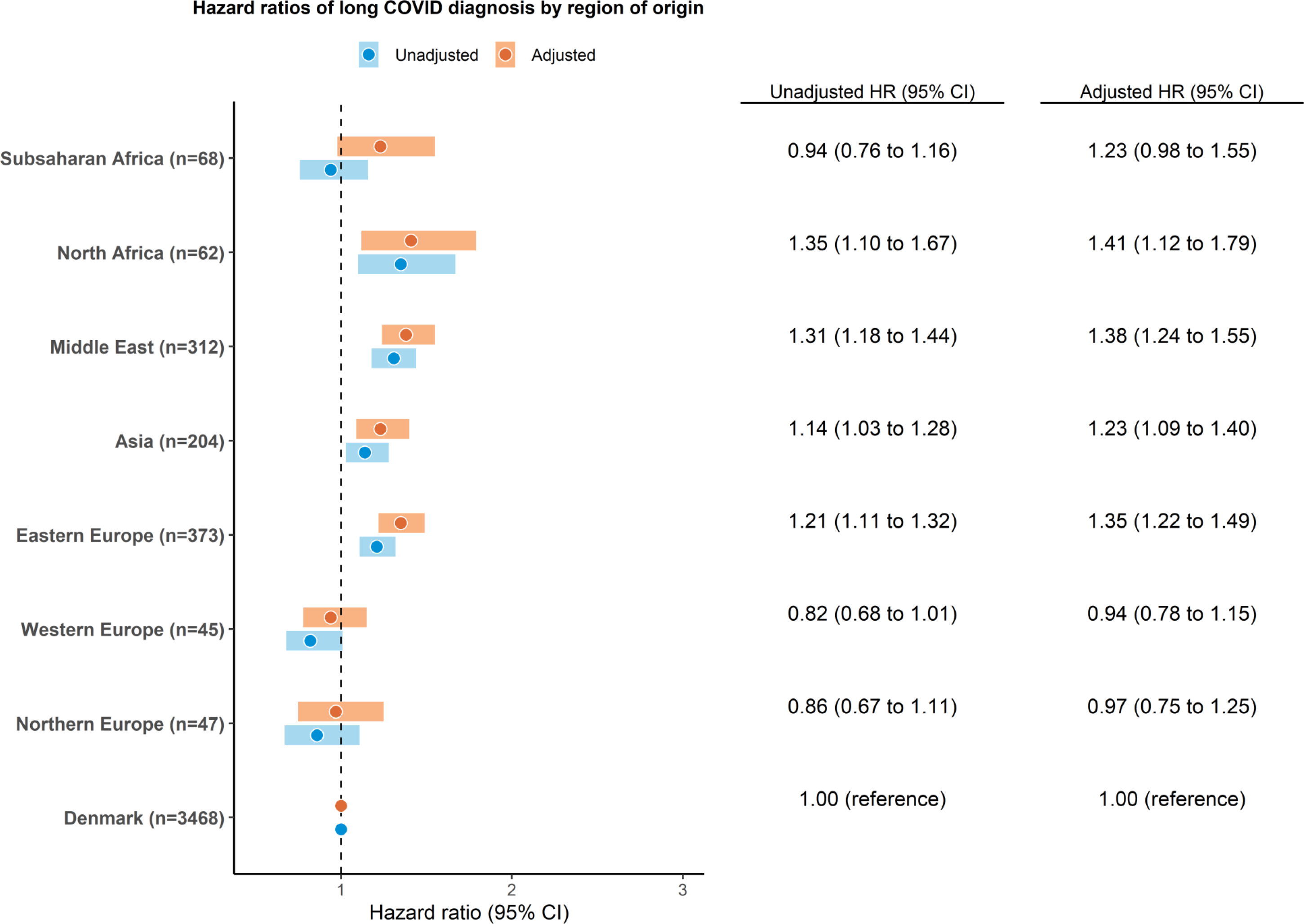
Hazard ratios of long COVID diagnosis by region of origin. The adjusted model composed age, sex, civil status, education, family income, and Charlson comorbidity index. HR=hazard ratio. CI=confidence interval.

**Figure 3.**
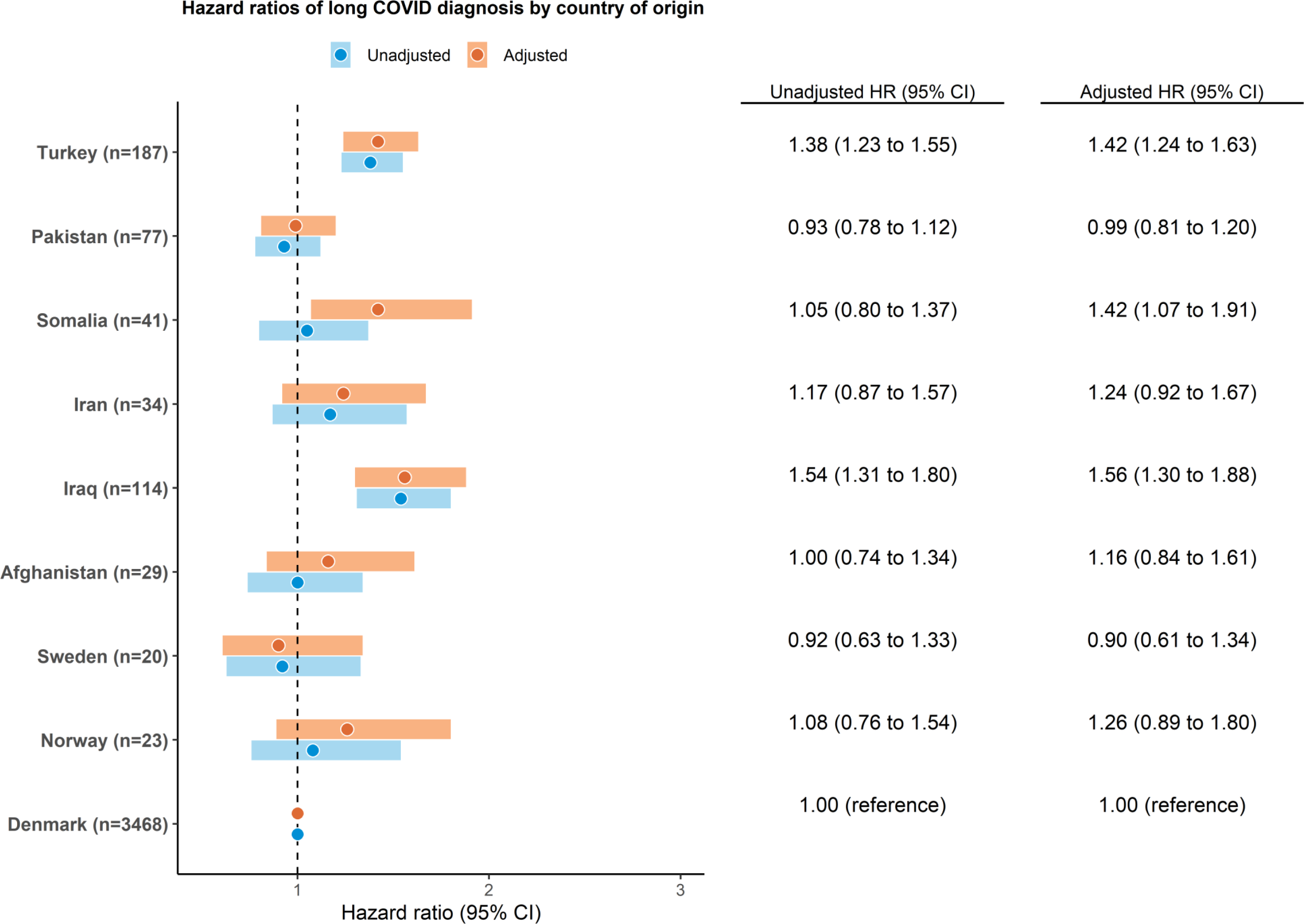
Hazard ratios of long COVID diagnosis by largest countries of origin. The adjusted model composed age, sex, civil status, education, family income, and Charlson comorbidity index. HR=hazard ratio. CI=confidence interval.

When investigating factors associated with increased risk of long COVID diagnosis, we observed that of all ethnic-age groups, the risk of long COVID diagnosis was highest among Subsaharan African aged >60 years (aHR 1.72; 95% CI 1.17–2.52) (Supplementary Table S3). Compared with native Danes non-hospitalised, COVID-19 hospitalisation was significantly associated with a higher risk of long COVID diagnosis among both native Danes and ethnic minorities before and after adjustment for confounders (Table 2). However, the hazard ratios of long COVID diagnosis for ethnic minorities like North African (aHR 3.98; 95% CI 2.75–5.75), Middle Eastern (aHR 4.43; 95% CI 3.71–5.29), Eastern European (aHR 4.49; 95% CI 3.84–5.23), Asian (aHR 3.44; 95% CI 2.79–4.23), and Subsaharan African (aHR 4.30; 95% CI 3.05–6.07) were still higher than that of native Danes (aHR 2.82; 95% CI 2.64–3.00) among individuals hospitalised for COVID-19. Further analysis showed that individuals not receiving COVID-19 vaccine exhibited greater risk of long COVID diagnosis than individuals vaccinated; and the association was found in native Danes only (aHR 1.47; 95% CI 1.33–1.63).

**Table 2.** Hazard ratios of long COVID diagnosis by hospitalisation for COVID-19.

|  | **COVID-19**<br>**hospitalisation** | **n** | **Unadjusted**<br>**HR (95% CI)** | **Adjusted**<br>**HR (95% CI)** |
| --- | --- | --- | --- | --- |
| Denmark | No | 1985 | 1.00 (reference) | 1.00 (reference) |
|  | Yes | 1483 | 6.53 (6.14 to 6.95) | 2.82 (2.64 to 3.00) |
| Northern Europe | No | 25 | 0.79 (0.58 to 1.07) | 0.90 (0.66 to 1.23) |
|  | Yes | 22 | 7.58 (4.98 to 11.52) | 3.44 (2.21 to 5.34) |
| Western Europe | No | 22 | 0.59 (0.43 to 0.81) | 0.73 (0.53 to 1.01) |
|  | Yes | 23 | 6.15 (4.08 to 9.28) | 2.57 (1.69 to 3.92) |
| Eastern Europe | No | 169 | 1.05 (0.94 to 1.17) | 1.15 (1.02 to 1.30) |
|  | Yes | 204 | 10.67 (9.27 to 12.29) | 4.49 (3.84 to 5.23) |
| Asia | No | 96 | 1.04 (0.90 to 1.20) | 1.14 (0.98 to 1.33) |
|  | Yes | 108 | 8.83 (7.29 to 10.70) | 3.44 (2.79 to 4.23) |
| Middle East | No | 151 | 1.14 (1.01 to 1.29) | 1.21 (1.05 to 1.39) |
|  | Yes | 161 | 10.16 (8.67 to 11.90) | 4.43 (3.71 to 5.29) |
| North Africa | No | 29 | 1.25 (0.96 to 1.63) | 1.27 (0.94 to 1.71) |
|  | Yes | 33 | 8.31 (5.89 to 11.73) | 3.98 (2.75 to 5.75) |
| Subsaharan Africa | No | 31 | 0.77 (0.58 to 1.01) | 0.99 (0.73 to 1.33) |
|  | Yes | 37 | 8.48 (6.13 to 11.73) | 4.30 (3.05 to 6.07) |

The adjusted model composed age, sex, civil status, education, family income, and Charlson comorbidity index. HR=hazard ratio. CI=confidence interval.

### Hospital contacts related to long COVID symptoms

The majority of ethnic minority groups and native Danes exhibited higher probabilities and higher odds of hospital contacts related to fatigue, headache, cardiopulmonary symptoms, and *any* long COVID symptoms within 6 months after COVID-19 diagnosis as compared to 6 months before COVID-19 diagnosis in the adjusted estimates (Figure 4 and Supplementary Table S4). However, compared with native Danes, differences in odds ratios of hospital contacts related to cardiopulmonary symptoms and *any* long COVID symptoms were more pronounced among North African, Middle Eastern, Eastern European, Asian, and Northern European, especially beyond 4 weeks to 6 months after COVID-19 diagnosis in both unadjusted and adjusted estimates (Table 3). Although the Subsaharan African group did not show significant difference from native Danes in the odds of hospital contacts related to *any* long COVID symptoms, this group was observed to have higher odds of hospital contacts related to symptoms like fatigue, headache, and cardiopulmonary symptoms beyond 4 weeks to 6 months after COVID-19 diagnosis. Moreover, analysis by largest countries of origin revealed that ethnic minority groups, especially Swedes, Afghans, Iraqis, Iranians, Somalis, Pakistanis, and Turks had higher odds of hospital contacts related to *any* long COVID symptoms than native Danes, particularly beyond 4 weeks to 6 months after COVID-19 diagnosis in both unadjusted and adjusted models (Supplementary Tables S5 and S6).

**Figure 4.**
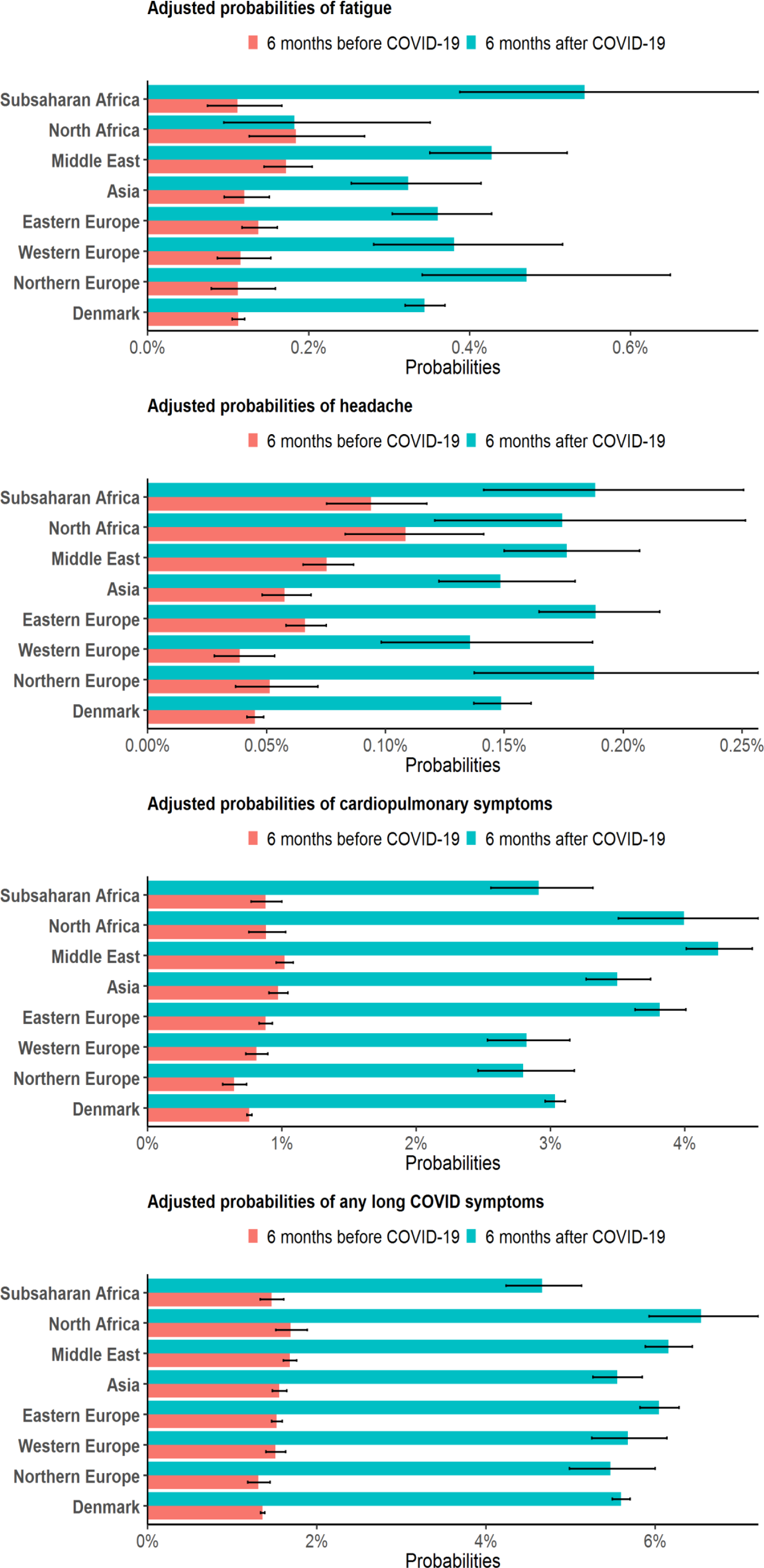
Adjusted probabilities of hospital contacts related to specific symptoms within 6 months after COVID-19 diagnosis compared with 6 months before COVID-19 diagnosis by region of origin. Hospital contacts related to cardiopulmonary symptoms included dyspnoea (difficulty in breathing), cough, and chest pain as a composite outcome. Hospital contacts related to any long COVID symptoms included fatigue, headache, dyspnoea (difficulty in breathing), cough, chest pain, depression and/or anxiety as a composite outcome. The adjusted model composed age, sex, civil status, education, family income, and Charlson comorbidity index.

**Table 3.**
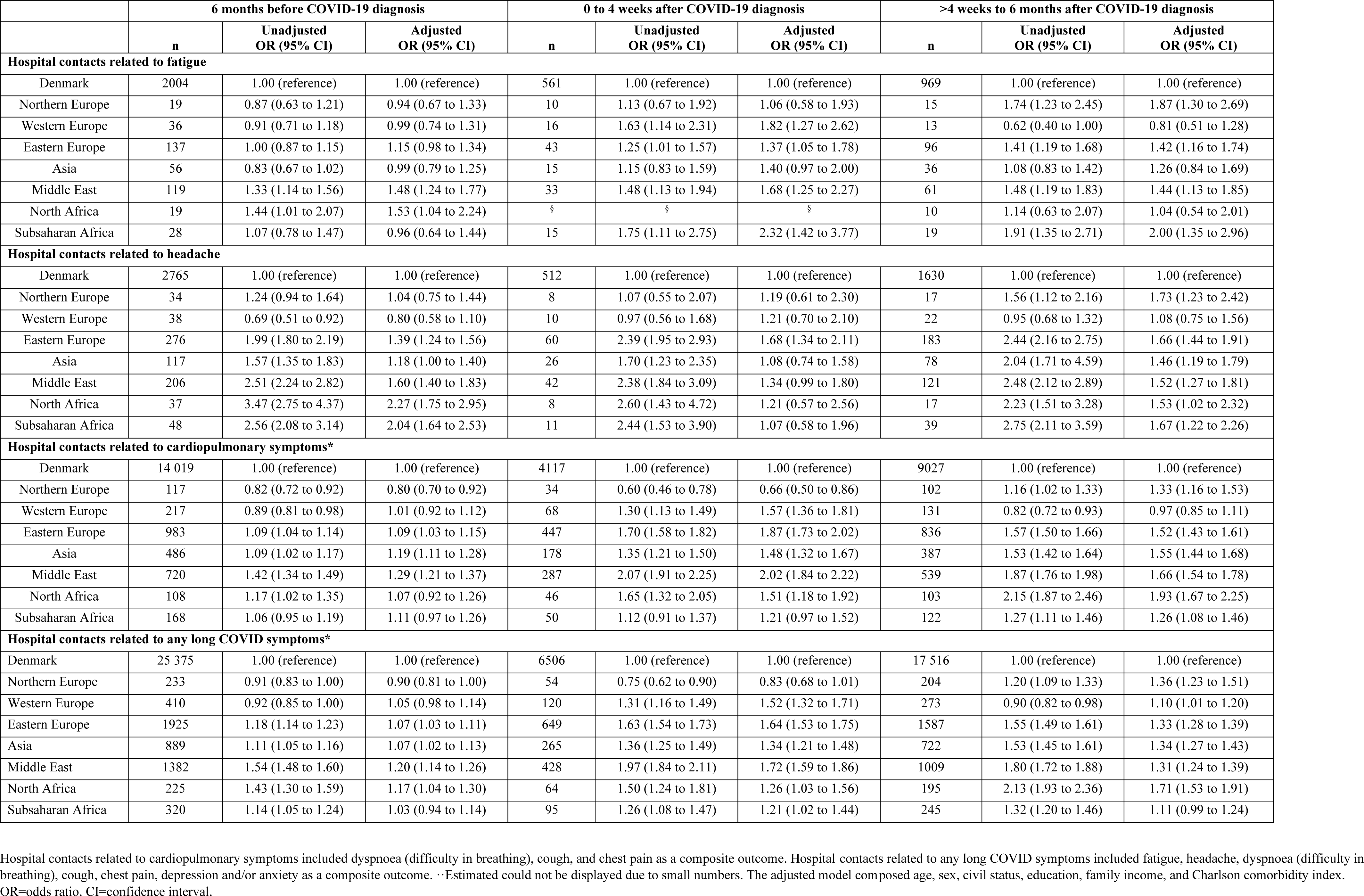
Odds ratios of hospital contacts related to specific symptoms by region of origin.

|  | 6 months before COVID-19 diagnosis |  |  | 0 to 4 weeks after COVID-19 diagnosis |  |  | >4 weeks to 6 months after COVID-19 diagnosis |  |  |
| --- | --- | --- | --- | --- | --- | --- | --- | --- | --- |
|  | n | Unadjusted<br>OR (95% CI) | Adjusted<br>OR (95% CI) | n | Unadjusted<br>OR (95% CI) | Adjusted<br>OR (95% CI) | n | Unadjusted<br>OR (95% CI) | Adjusted<br>OR (95% CI) |
| Hospital contacts related to fatigue |  |  |  |  |  |  |  |  |  |
| Denmark | 2004 | 1.00 (reference) | 1.00 (reference) | 561 | 1.00 (reference) | 1.00 (reference) | 969 | 1.00 (reference) | 1.00 (reference) |
| Northern Europe | 19 | 0.87 (0.63 to 1.21) | 0.94 (0.67 to 1.33) | 10 | 1.13 (0.67 to 1.92) | 1.06 (0.58 to 1.93) | 15 | 1.74 (1.23 to 2.45) | 1.87 (1.30 to 2.69) |
| Western Europe | 36 | 0.91 (0.71 to 1.18) | 0.99 (0.74 to 1.31) | 16 | 1.63 (1.14 to 2.31) | 1.82 (1.27 to 2.62) | 13 | 0.62 (0.40 to 1.00) | 0.81 (0.51 to 1.28) |
| Eastern Europe | 137 | 1.00 (0.87 to 1.15) | 1.15 (0.98 to 1.34) | 43 | 1.25 (1.01 to 1.57) | 1.37 (1.05 to 1.78) | 96 | 1.41 (1.19 to 1.68) | 1.42 (1.16 to 1.74) |
| Asia | 56 | 0.83 (0.67 to 1.02) | 0.99 (0.79 to 1.25) | 15 | 1.15 (0.83 to 1.59) | 1.40 (0.97 to 2.00) | 36 | 1.08 (0.83 to 1.42) | 1.26 (0.84 to 1.69) |
| Middle East | 119 | 1.33 (1.14 to 1.56) | 1.48 (1.24 to 1.77) | 33 | 1.48 (1.13 to 1.94) | 1.68 (1.25 to 2.27) | 61 | 1.48 (1.19 to 1.83) | 1.44 (1.13 to 1.85) |
| North Africa | 19 | 1.44 (1.01 to 2.07) | 1.53 (1.04 to 2.24) | § | § | § | 10 | 1.14 (0.63 to 2.07) | 1.04 (0.54 to 2.01) |
| Subsaharan Africa | 28 | 1.07 (0.78 to 1.47) | 0.96 (0.64 to 1.44) | 15 | 1.75 (1.11 to 2.75) | 2.32 (1.42 to 3.77) | 19 | 1.91 (1.35 to 2.71) | 2.00 (1.35 to 2.96) |
| Hospital contacts related to headache |  |  |  |  |  |  |  |  |  |
| Denmark | 2765 | 1.00 (reference) | 1.00 (reference) | 512 | 1.00 (reference) | 1.00 (reference) | 1630 | 1.00 (reference) | 1.00 (reference) |
| Northern Europe | 34 | 1.24 (0.94 to 1.64) | 1.04 (0.75 to 1.44) | 8 | 1.07 (0.55 to 2.07) | 1.19 (0.61 to 2.30) | 17 | 1.56 (1.12 to 2.16) | 1.73 (1.23 to 2.42) |
| Western Europe | 38 | 0.69 (0.51 to 0.92) | 0.80 (0.58 to 1.10) | 10 | 0.97 (0.56 to 1.68) | 1.21 (0.70 to 2.10) | 22 | 0.95 (0.68 to 1.32) | 1.08 (0.75 to 1.56) |
| Eastern Europe | 276 | 1.99 (1.80 to 2.19) | 1.39 (1.24 to 1.56) | 60 | 2.39 (1.95 to 2.93) | 1.68 (1.34 to 2.11) | 183 | 2.44 (2.16 to 2.75) | 1.66 (1.44 to 1.91) |
| Asia | 117 | 1.57 (1.35 to 1.83) | 1.18 (1.00 to 1.40) | 26 | 1.70 (1.23 to 2.35) | 1.08 (0.74 to 1.58) | 78 | 2.04 (1.71 to 4.59) | 1.46 (1.19 to 1.79) |
| Middle East | 206 | 2.51 (2.24 to 2.82) | 1.60 (1.40 to 1.83) | 42 | 2.38 (1.84 to 3.09) | 1.34 (0.99 to 1.80) | 121 | 2.48 (2.12 to 2.89) | 1.52 (1.27 to 1.81) |
| North Africa | 37 | 3.47 (2.75 to 4.37) | 2.27 (1.75 to 2.95) | 8 | 2.60 (1.43 to 4.72) | 1.21 (0.57 to 2.56) | 17 | 2.23 (1.51 to 3.28) | 1.53 (1.02 to 2.32) |
| Subsaharan Africa | 48 | 2.56 (2.08 to 3.14) | 2.04 (1.64 to 2.53) | 11 | 2.44 (1.53 to 3.90) | 1.07 (0.58 to 1.96) | 39 | 2.75 (2.11 to 3.59) | 1.67 (1.22 to 2.26) |
| Hospital contacts related to cardiopulmonary symptoms* |  |  |  |  |  |  |  |  |  |
| Denmark | 14 019 | 1.00 (reference) | 1.00 (reference) | 4117 | 1.00 (reference) | 1.00 (reference) | 9027 | 1.00 (reference) | 1.00 (reference) |
| Northern Europe | 117 | 0.82 (0.72 to 0.92) | 0.80 (0.70 to 0.92) | 34 | 0.60 (0.46 to 0.78) | 0.66 (0.50 to 0.86) | 102 | 1.16 (1.02 to 1.33) | 1.33 (1.16 to 1.53) |
| Western Europe | 217 | 0.89 (0.81 to 0.98) | 1.01 (0.92 to 1.12) | 68 | 1.30 (1.13 to 1.49) | 1.57 (1.36 to 1.81) | 131 | 0.82 (0.72 to 0.93) | 0.97 (0.85 to 1.11) |
| Eastern Europe | 983 | 1.09 (1.04 to 1.14) | 1.09 (1.03 to 1.15) | 447 | 1.70 (1.58 to 1.82) | 1.87 (1.73 to 2.02) | 836 | 1.57 (1.50 to 1.66) | 1.52 (1.43 to 1.61) |
| Asia | 486 | 1.09 (1.02 to 1.17) | 1.19 (1.11 to 1.28) | 178 | 1.35 (1.21 to 1.50) | 1.48 (1.32 to 1.67) | 387 | 1.53 (1.42 to 1.64) | 1.55 (1.44 to 1.68) |
| Middle East | 720 | 1.42 (1.34 to 1.49) | 1.29 (1.21 to 1.37) | 287 | 2.07 (1.91 to 2.25) | 2.02 (1.84 to 2.22) | 539 | 1.87 (1.76 to 1.98) | 1.66 (1.54 to 1.78) |
| North Africa | 108 | 1.17 (1.02 to 1.35) | 1.07 (0.92 to 1.26) | 46 | 1.65 (1.32 to 2.05) | 1.51 (1.18 to 1.92) | 103 | 2.15 (1.87 to 2.46) | 1.93 (1.67 to 2.25) |
| Subsaharan Africa | 168 | 1.06 (0.95 to 1.19) | 1.11 (0.97 to 1.26) | 50 | 1.12 (0.91 to 1.37) | 1.21 (0.97 to 1.52) | 122 | 1.27 (1.11 to 1.46) | 1.26 (1.08 to 1.46) |
| Hospital contacts related to any long COVID symptoms* |  |  |  |  |  |  |  |  |  |
| Denmark | 25 375 | 1.00 (reference) | 1.00 (reference) | 6506 | 1.00 (reference) | 1.00 (reference) | 17 516 | 1.00 (reference) | 1.00 (reference) |
| Northern Europe | 233 | 0.91 (0.83 to 1.00) | 0.90 (0.81 to 1.00) | 54 | 0.75 (0.62 to 0.90) | 0.83 (0.68 to 1.01) | 204 | 1.20 (1.09 to 1.33) | 1.36 (1.23 to 1.51) |
| Western Europe | 410 | 0.92 (0.85 to 1.00) | 1.05 (0.98 to 1.14) | 120 | 1.31 (1.16 to 1.49) | 1.52 (1.32 to 1.71) | 273 | 0.90 (0.82 to 0.98) | 1.10 (1.01 to 1.20) |
| Eastern Europe | 1925 | 1.18 (1.14 to 1.23) | 1.07 (1.03 to 1.11) | 649 | 1.63 (1.54 to 1.73) | 1.64 (1.53 to 1.75) | 1587 | 1.55 (1.49 to 1.61) | 1.33 (1.28 to 1.39) |
| Asia | 889 | 1.11 (1.05 to 1.16) | 1.07 (1.02 to 1.13) | 265 | 1.36 (1.25 to 1.49) | 1.34 (1.21 to 1.48) | 722 | 1.53 (1.45 to 1.61) | 1.34 (1.27 to 1.43) |
| Middle East | 1382 | 1.54 (1.48 to 1.60) | 1.20 (1.14 to 1.26) | 428 | 1.97 (1.84 to 2.11) | 1.72 (1.59 to 1.86) | 1009 | 1.80 (1.72 to 1.88) | 1.31 (1.24 to 1.39) |
| North Africa | 225 | 1.43 (1.30 to 1.59) | 1.17 (1.04 to 1.30) | 64 | 1.50 (1.24 to 1.81) | 1.26 (1.03 to 1.56) | 195 | 2.13 (1.93 to 2.36) | 1.71 (1.53 to 1.91) |
| Subsaharan Africa | 320 | 1.14 (1.05 to 1.24) | 1.03 (0.94 to 1.14) | 95 | 1.26 (1.08 to 1.47) | 1.21 (1.02 to 1.44) | 245 | 1.32 (1.20 to 1.46) | 1.11 (0.99 to 1.24) |

Hospital contacts related to cardiopulmonary symptoms included dyspnoea (difficulty in breathing), cough, and chest pain as a composite outcome. Hospital contacts related to any long COVID symptoms included fatigue, headache, dyspnoea (difficulty in breathing), cough, chest pain, depression and/or anxiety as a composite outcome. ·-Estimated could not be displayed due to small numbers. The adjusted model composed age, sex, civil status, education, family income, and Charlson comorbidity index. OR=odds ratio. CI=confidence interval.

## Discussion

This Danish nationwide cohort study found that compared with native Danes, the risk of long COVID diagnosis was higher in the majority of ethnic minorities, notably for North African, Middle Eastern, Eastern European, and Asian in both unadjusted and adjusted models. Our findings also confirm that the chances of reporting cardiopulmonary symptoms (including dyspnoea, cough, and chest pain) and *any* long COVID symptoms were higher among North African, Middle Eastern, Eastern European, and Asian than among native Danes, especially beyond 4 weeks to 6 months after COVID-19 diagnosis in both unadjusted and adjusted models. In the analysis by largest countries of origin, this study found that the risk of long COVID diagnosis was higher in Iraqis, Turks, and Somalis than in native Danes after adjustment for all relevant covariates. While chances of reporting *any* long COVID symptoms were higher in Swedes, Afghans, Iraqis, Iranians, Somalis, Pakistanis, and Turks than in native Danes, particularly beyond 4 weeks to 6 months after COVID-19 diagnosis in both unadjusted and adjusted estimates.

The observed higher risk of long COVID among ethnic minority groups than native Danes may be explained by several factors including older age, COVID-19 hospitalisation, use of intensive care, and comorbidities as previously reported [2,3]. Compared with native Danes, older age (>60 years) was only observed as a contributing factor to the increased risk of long COVID in the Subsaharan African group. Another factor that was found to contribute to the risk of long COVID was COVID-19 hospitalisation (a marker of disease severity). Our estimates demonstrate that COVID-19 hospitalisation was associated with increased risk of long COVID in both ethnic minorities and native Danes. Notwithstanding the increased risk of long COVID in native Danes hospitalised for COVID-19, the present study found that ethnic minorities were more likely than native Danes to be hospitalised for COVID-19. In addition, differences in hazard ratios of long COVID diagnosis for North African, Middle Eastern, Eastern European, Asian, and Subsaharan African compared with that of native Danes hospitalised for COVID-19 were substantial. Hence, this may signify a greater burden of long COVID in these ethnic minority groups. Furthermore, the use of intensive care may partly contribute to the increased risk of long COVID among ethnic minorities. A recent Danish study using data from patients hospitalised for COVID-19 has reported that ethnic minorities originating from non-Western countries had a higher chance than native Danes of use of mechanical ventilation [29], which could be seen as another marker of COVID-19 severity associated with high burden of long COVID found in ethnic minorities living in Denmark. Apart from markers of disease severity, the risk of long COVID may also be influenced by presence of comorbidities. Our estimates show that individuals originating from North Africa and Middle East as well as those from Pakistan, Turkey, Iraq, Iran, and Afghanistan were more likely than native Danes to have a high burden of comorbidities.

Despite considering a wide range of comorbidities using Charlson comorbidity index in our models, the risk of long COVID remained significantly higher in ethnic minority groups reported. Therefore, this work suggests that the high burden of long COVID in this population may also be rooted in the complex interplay between the COVID-19 variant, immunological factors, and circulatory system [30]. In line with our findings, previous studies in the United States, United Kingdom, and Netherlands have reported that individuals of African, Asian, and Turkish origin exhibited higher chances of reporting long COVID symptoms than the native majority population [14]. Recent evidence also shows that individuals not receiving COVID-19 vaccine have a higher risk of long COVID in the general population.^15^ Although the majority of ethnic minorities were less likely than native Danes to receive COVID-19 vaccine, their risk of long COVID did not differ by COVID-19 vaccination, indicating other factors such as barriers to accessing healthcare, differences in healthcare demand, and late contact with healthcare when having COVID-19 may have contributed to their increased risk of long COVID.

### Strengths and limitations

Compared with previous research, the present study has several strengths, including using a nationwide sample of individuals diagnosed with COVID-19 in Denmark, using an established ICD-10 based diagnosis of long COVID in Denmark, and incorporating a wide range of comorbidities and sociodemographic factors. To our knowledge, no previous study on long COVID in ethnic minorities that had included symptoms experienced before COVID-19 diagnosis. Overall, studies on long COVID among ethnic minorities are still lacking and this study is among the few to contribute knowledge to the existing body of literature. However, there are some limitations. First, long COVID diagnosis in the registers was implemented from April 2020, which entails lack of registration of long COVID cases between January and March 2020 [26]. Second, symptoms included in the study were in connection with hospital contact and identified by ICD-10 codes, which may have introduced some selection and registration bias. This is because these symptoms may not be representative of long COVID situation in Denmark as some may choose not to contact hospital if the symptoms are not interfering with their daily routines. Additionally, the registration of symptoms might also be problematic as individuals are more likely to receive clinical-based diagnoses rather than symptoms-based diagnoses in hospital setting. For that reason, we could not perform analysis of a single symptom by largest countries of origin due to small sample size. Therefore, it is possible that individuals are experiencing long COVID symptoms more than what it is reported. We were unable to account for health-seeking behaviours and cultural norms associated with healthcare contact as such data were unavailable. It may be the case that some ethnic minority groups have a certain tendency of contacting the hospital, which may have influenced our estimates.

Our results have implications for clinical work and research. First, these findings raise intriguing question regarding preparedness and resilience of healthcare system in highly anticipated burden of long COVID. This implies that the health care system that dealt with the acute consequences of COVID-19 now also has to consider a new situation of long-term and indirect consequences of the pandemic which puts new demands on health care professionals and the society. Second, the higher risk of long COVID in ethnic minorities is a major concern for equity in health and addressing this health problem may require multisectoral response, funding, care, and treatment approaches which are culturally acceptable to this population. Lastly, future research should also focus on understanding key drivers of long COVID and impact of long COVID on sick-leave and labour market participation among ethnic minorities.

## Ethics statement

This study was approved by the Danish Data Protection Agency, reference number 514-0670/21-3000. No further approval is required regarding registry-based research.

## Data availability

Data that supports the findings of this work are stored at Statistics Denmark and are not publicly available. Data access may be granted upon approval from the relevant data custodians.

## Acknowledgements

None

## Authors’ contributions

GFM, CA, TB, MR, AC, JHP, and MN conceived the study and analytical strategy. GFM performed the literature review, analysed cohort data and prepared results. GFM, CA, TB, MR, AC, JHP, and MN contributed to the interpretation of results. GFM wrote the first draft of the report. All authors commented on the report draft and approved the final text.

## Supplementary material

### Search strategy for literature review

#### Block 1

“Post-Acute COVID-19 Syndrome” OR “Post-Acute COVID-19 Syndromes” OR “COVID-19 Syndrome, Post-Acute” OR “Long Haul COVID-19” OR “COVID-19, Long Haul” OR “Long Haul COVID 19” OR “Long Haul COVID-19s” OR “Post Acute COVID-19 Syndrome” OR “Post Acute COVID 19 Syndrome” OR “Long COVID” OR “Post-Acute Sequelae of SARS-CoV-2 Infection” OR “Post Acute Sequelae of SARS CoV 2 Infection” OR “Post COVID Conditions” OR “Post-COVID Conditions” OR “Post-COVID Condition” OR “Long-Haul COVID” OR “COVID, Long-Haul” OR “Long Haul COVID” OR “Long-Haul COVIDs”

AND

#### Block 2

“migrant*” OR “transient” OR “refugee*” OR “asylum seeker*” OR “displaced person*” OR “asylee” OR “immigrant*” OR “foreigner*” OR “emigrant*” OR “ethnic minority” OR “racial minority” OR “diaspora*” OR “human migration” OR “undocumented immigrant*” OR “undocumented worker*” OR “unauthorized immigrant*” OR “Black American*” OR “African American*” OR “black” OR “Afro-American*” or “African*” OR “Hispanic” OR “Latino” or “Latinx” OR “Hispanic*” OR “Latin American*” OR “Asian” OR “Asian American” OR “Middle Eastern” OR “Southwestern Asian” OR “ethnicity” OR “ethnic group” OR “nationality” OR “race” OR “racial group*” OR “Continental Population Group” OR “BAME” OR “BIPOC” OR “native-born*” or “indigenous people*” or “native people*”

**Table S1.** ICD-10 codes of symptoms and diagnoses.

| **Symptom** | **ICD-10 codes** |
| --- | --- |
| Fatigue | R53, R539, R539A, R539C, R539E, R539F, G933 |
| Headache | R51–R519 |
| Dyspnoea (difficulty in breathing) | R06, R060, R060A, R061, R063, R064, R065, R066, R068, R068A, R068B, R068C, R068D, R068E |
| Cough | R05–R059 |
| Chest pain | R07–R074 |
| Depression/anxiety | F32–F339, F411–F418, F430–F4300 |
| **Diagnosis** |  |
| COVID-19 | B34.2, B34.2A, B97.2, B97.2A |
| Long COVID | B94.8, B94.8A |
| Myocardial infarction | I21–I21.9, I22, I25.2 |
| Congestive heart failure | I10–I10.9, I11, I11.0, I13.0, I13.2, I25.5, I42.0, I42.6, I42.7, I42.8, I42.9, I43, I50, I50.0, I50.1, I50.9 |
| Peripheral vascular disease | I70, I71, I73.1, I73.8, I73.9, I77.1, I79.0, I79.2 |
| Cerebrovascular disease | G45, I60–I64, I67, I69 |
| Chronic obstructive pulmonary disease | J43, J44–J44.9 |
| Rheumatic disease | M05, M06, M12.3, M07.0–M07.3 |
| Dementia | F00–F03, F05.1, G30, G31.1, G31.9 |
| Peptic ulcer disease | K25–K28 |
| Hemiplegia | G11.4, G80, G81, G82, G83.0–G83.3, G83.8 |
| Diabetes without complications | E10, E11, E10.0, E10.0, E10.1, E11.0, E11.1, E12.0, E12.1, E13.0, E13.1, E14.0, E14.1 |
| Diabetes with complications | E10, E11, E10.2, E10.5, E10.7, E11.2, E11.7, E12.2, E12.7, E13.2, E13.7, E14.2, E14.7 |
| Mild liver disease | B15–B19, K70, K70.0, K70.1 |
| Moderate to severe liver disease | I85.0, I85.9, I98.2, I98.3, K70.3, K70.4, K70.9, K73, K74.0, K74.6, K75.4 |
| Renal disease | I12.0, I13.1, N03.2–N03.7, N05.2–N05.7, N11, N18, N19, N25.0, Q61.1, Z49, Z94.0, Z99.2 |
| Malignancy | C00–C09, C10–C41, C45–C58, C60–C76, C81–C86, C88–C97 |
| Metastatic cancer | C77–C80 |
| Acquired immunodeficiency syndrome | B20–B24, R75, Z21 |

**Figure S1.**
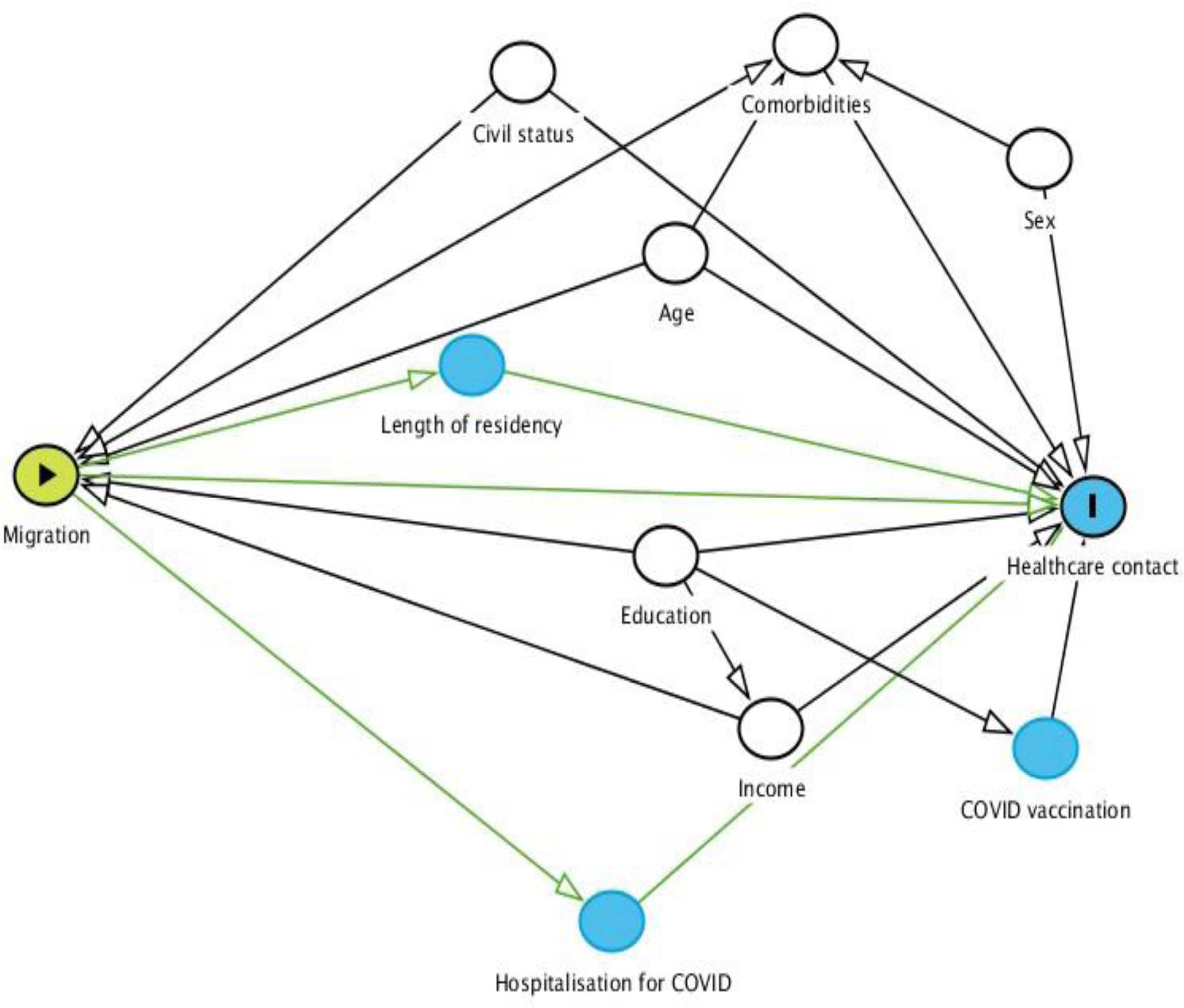
Directed Acyclic Graphs for confounders assessment. Age, sex, civil status, comorbidities, education, and income were identified as confounders.

**Table S2.** Individuals who had first-time tested positive for SARS-COV-2 between January 2020 and August 2022 by largest countries of origin.

|  | Denmark | Norway | Sweden | Afghanistan | Iraq | Iran | Somalia | Pakistan | Turkey |
| --- | --- | --- | --- | --- | --- | --- | --- | --- | --- |
| n | 1 952 021 | 7200 | 7078 | 9273 | 15 600 | 9200 | 9077 | 12 462 | 35 460 |
| Immigrants | NA | 6680 (92.8%) | 6395 (90.4%) | 8455 (91.2%) | 12 571 (80.1%) | 7667 (83.3%) | 6274 (69.1%) | 7031 (56.4%) | 18 764 (52.9%) |
| Descendants | NA | 520 (7.2%) | 683 (9.6%) | 818 (8.8%) | 3119 (19.9%) | 1533 (16.7%) | 2803 (30.9%) | 5431 (43.6%) | 16 696 (47.1%) |
| Length of residency, years | NA | 36 (17–41) | 36 (16–38) | 19 (16–21) | 22 (19–26) | 27 (14–34) | 24 (20–27) | 36 (29–39) | 34 (26–38) |
| Age, years | 61 (43–75) | 62 (39–76) | 64 (41–76) | 44 (32–58) | 48 (32–60) | 49 (36–60) | 43 (28–56) | 56 (42–71) | 49 (36–62) |
| Sex |  |  |  |  |  |  |  |  |  |
| Female | 1 026 373 (52.6%) | 4752 (66.0%) | 4308 (60.8%) | 4470 (48.2%) | 7807 (49.8%) | 4400 (47.8%) | 4856 (53.4%) | 6415 (51.5%) | 18 358 (51.8%) |
| Male | 925 648 (47.4%) | 2448 (34.0%) | 2770 (39.2%) | 4803 (51.8%) | 7793 (50.2%) | 4800 (52.2%) | 4221 (46.6%) | 6047 (48.5%) | 17 102 (48.2%) |
| Civil status |  |  |  |  |  |  |  |  |  |
| Cohabiting | 862 240 (44.2%) | 2615 (36.3%) | 2903 (41.0%) | 4211 (45.4%) | 6402 (40.8%) | 4126 (44.8%) | 2030 (22.4%) | 7832 (62.8%) | 19 954 (56.3%) |
| Living alone | 852 792 (43.7%) | 3824 (53.1%) | 3464 (48.9%) | 4610 (49.7%) | 7730 (49.3%) | 3832 (41.7%) | 5894 (64.9%) | 3672 (29.5%) | 11 723 (33.0%) |
| Other | 236 989 (12.1%) | 761 (10.6%) | 711 (10.1%) | 452 (4.9%) | 1558 (9.9%) | 1242 (13.5%) | 1153 (12.7%) | 958 (7.7%) | 3783 (10.7%) |
| Education |  |  |  |  |  |  |  |  |  |
| Low | 465 063 (23.8%) | 597 (8.3%) | 663 (9.4%) | 3363 (36.3%) | 6399 (40.8%) | 2365 (25.7%) | 4684 (51.6%) | 4426 (35.5%) | 15 902 (44.8%) |
| Medium | 904 288 (46.3%) | 2464 (34.2%) | 2406 (34.0%) | 3183 (34.3%) | 5242 (33.4%) | 3060 (33.3%) | 2724 (30.0%) | 4086 (32.8%) | 11 904 (33.6%) |
| High | 571 393 (29.3%) | 3769 (52.4%) | 3522 (49.7%) | 1581 (17.0%) | 2728 (17.4%) | 3309 (36.0%) | 682 (7.5%) | 3293 (26.4%) | 5304 (15.0%) |
| Missing | 11 277 (0.6%) | 370 (5.1%) | 487 (6.9%) | 1146 (12.4%) | 1321 (8.4%) | 466 (5.0%) | 987 (10.9%) | 657 (5.3%) | 2350 (6.6%) |
| Family income |  |  |  |  |  |  |  |  |  |
| Low | 353 452 (18.1%) | 2441 (33.9%) | 1877 (26.5%) | 5783 (62.4%) | 9801 (62.5%) | 4073 (44.3%) | 6921 (76.3%) | 6437 (51.7%) | 16 345 (46.1%) |
| Middle | 585 134 (30.0%) | 1423 (19.8%) | 1541 (21.8%) | 1896 (20.4%) | 2845 (18.1%) | 2042 (22.2%) | 1108 (12.2%) | 3243 (26.0%) | 11 055 (31.2%) |
| High | 859 757 (44.0%) | 2728 (37.9%) | 3160 (44.6%) | 839 (9.1%) | 1571 (10.0%) | 2279 (24.8%) | 249 (2.7%) | 1904 (15.3%) | 5632 (15.9%) |
| Missing | 153 678 (7.9%) | 608 (8.4%) | 500 (7.1%) | 755 (8.1%) | 1473 (9.4%) | 806 (8.7%) | 799 (8.8%) | 878 (7.0%) | 2428 (6.8%) |
| COVID-19 hospitalisation | 30 230 (1.5%) | 123 (1.7%) | 142 (2.0%) | 370 (3.9%) | 831 (5.1%) | 366 (3.9%) | 306 (3.3%) | 618 (4.8%) | 1547 (4.3%) |
| Intensive care | 12 014 (0.6%) | 35 (0.5%) | 37 (0.5%) | 28 (0.3%) | 78 (0.5%) | 36 (0.4%) | 68 (0.7%) | 74 (0.6%) | 160 (0.4%) |
| COVID-19 vaccination |  |  |  |  |  |  |  |  |  |
| One dose | 1 813 312 (92.9%) | 6320 (87.8%) | 6258 (88.4%) | 7565 (81.5%) | 11 037 (70.3%) | 7859 (85.4%) | 5713 (62.9%) | 9957 (79.9%) | 23 754 (67.0%) |
| Two doses | 1 796 381 (92.0%) | 6199 (86.1%) | 6184 (87.4%) | 7246 (78.1%) | 10 482 (66.8%) | 7689 (83.6%) | 5248 (57.8%) | 9586 (76.9%) | 22 559 (63.6%) |
| Three doses | 1 489 444 (76.3%) | 4889 (67.9%) | 4856 (68.6%) | 3667 (39.5%) | 4684 (29.8%) | 5278 (57.3%) | 1586 (17.5%) | 4523 (36.3%) | 10 760 (30.3%) |
| Charlson comorbidity index* |  |  |  |  |  |  |  |  |  |
| 0 | 1 550 412 (79.4%) | 5971 (82.9%) | 5789 (81.8%) | 6974 (75.2%) | 11 356 (72.4%) | 6774 (73.6%) | 7179 (79.1%) | 8900 (71.4%) | 25 568 (72.1%) |
| 1–2 | 396 200 (20.3%) | 1210 (16.8%) | 1274 (18.0%) | 2291 (24.7%) | 4318 (27.5%) | 2410 (26.2%) | 1884 (20.8%) | 3549 (28.5%) | 9856 (27.8%) |
| ≥3 | 5409 (0.3%) | 19 (0.3%) | 15 (0.2%) | 8 (0.1%) | 16 (0.1%) | 16 (0.2%) | 14 (0.1%) | 13 (0.1%) | 36 (0.1%) |

Data are in median (IQR) or n (%). \*Charlson comorbidity index composed myocardial infarction, congestive heart failure, peripheral vascular disease, cerebrovascular disease, chronic obstructive pulmonary disease, rheumatic disease, dementia, peptic ulcer disease, hemiplegia, diabetes without complications, diabetes with complications, mild liver disease, moderate to severe liver disease, renal disease, malignancy, metastatic cancer, and acquired immunodeficiency syndrome (AIDS). NA=not applicable.

**Table S3.** Hazard ratios of long COVID diagnosis by age group.

|  | Age group | n | Unadjusted<br>HR (95% CI) | Adjusted<br>HR (95% CI) |
| --- | --- | --- | --- | --- |
| Denmark | 18–60 | 2126 | 1.00 (reference) | 1.00 (reference) |
|  | >60 | 1342 | 0.76 (0.72 to 0.80) | 0.52 (0.49 to 0.55) |
| Northern Europe | 18–60 | 32 | 1.08 (0.79 to 1.48) | 1.44 (1.05 to 1.98) |
|  | >60 | 15 | 0.50 (0.33 to 0.75) | 0.34 (0.22 to 0.52) |
| Western Europe | 18–60 | 26 | 0.59 (0.42 to 0.83) | 0.74 (0.52 to 1.06) |
|  | >60 | 19 | 0.62 (0.43 to 0.90) | 0.47 (0.33 to 0.68) |
| Eastern Europe | 18–60 | 293 | 1.04 (0.93 to 1.15) | 1.19 (1.06 to 1.33) |
|  | >60 | 80 | 1.15 (0.97 to 1.35) | 0.85 (0.70 to 1.04) |
| Asia | 18–60 | 150 | 0.95 (0.83 to 1.10) | 0.93 (0.79 to 1.08) |
|  | >60 | 54 | 1.13 (0.92 to 1.38) | 1.11 (0.90 to 1.37) |
| Middle East | 18–60 | 243 | 1.10 (0.99 to 1.24) | 1.16 (1.02 to 1.32) |
|  | >60 | 69 | 1.31 (1.07 to 1.61) | 1.04 (0.83 to 1.32) |
| North Africa | 18–60 | 47 | 1.38 (1.08 to 1.77) | 1.38 (1.06 to 1.79) |
|  | >60 | 15 | 0.83 (0.55 to 1.26) | 0.60 (0.36 to 1.01) |
| Subsaharan Africa | 18–60 | 53 | 0.67 (0.52 to 0.86) | 0.85 (0.64 to 1.12) |
|  | >60 | 15 | 1.60 (1.11 to 2.31) | 1.72 (1.17 to 2.52) |

The adjusted model composed age, sex, civil status, education, family income, and Charlson comorbidity index. HR=hazard ratio. CI=confidence interval.

**Table S4.** Odds ratio of hospital contacts related to specific symptoms 6 months after COVID-19 diagnosis compared with 6 months before COVID-19 diagnosis by region of origin.

|  | 6 months before<br>COVID-19 diagnosis | 6 months after COVID-19 diagnosis |  |
| --- | --- | --- | --- |
|  | OR (95% CI) | Unadjusted<br>OR (95% CI) | Adjusted<br>OR (95% CI) |
| **Hospital contacts related to fatigue** |  |  |  |
| Denmark | 1.00 (reference) | 3.10 (2.96 to 3.25) | 3.06 (2.91 to 3.21) |
| Northern Europe | 1.00 (reference) | 4.41 (2.84 to 6.84) | 4.21 (2.64 to 6.71) |
| Western Europe | 1.00 (reference) | 2.93 (1.99 to 4.31) | 3.31 (2.20 to 4.97) |
| Eastern Europe | 1.00 (reference) | 2.76 (2.27 to 3.36) | 2.62 (2.10 to 3.27) |
| Asia | 1.00 (reference) | 2.62 (1.94 to 3.54) | 2.70 (1.94 to 3.76) |
| Middle East | 1.00 (reference) | 2.28 (1.81 to 2.87) | 2.49 (1.93 to 3.21) |
| North Africa | 1.00 (reference) | 1.07 (0.54 to 2.09) | 0.98 (0.46 to 2.10) |
| Subsaharan Africa | 1.00 (reference) | 2.95 (1.90 to 4.58) | 4.89 (2.90 to 8.24) |
| **Hospital contacts related to headache** |  |  |  |
| Denmark | 1.00 (reference) | 3.30 (3.16 to 3.45) | 3.30 (3.14 to 3.46) |
| Northern Europe | 1.00 (reference) | 3.05 (2.04 to 4.55) | 3.65 (2.34 to 5.70) |
| Western Europe | 1.00 (reference) | 3.91 (2.59 to 5.88) | 3.50 (2.24 to 5.46) |
| Eastern Europe | 1.00 (reference) | 2.63 (2.29 to 3.03) | 2.84 (2.43 to 3.33) |
| Asia | 1.00 (reference) | 2.65 (2.13 to 3.29) | 2.58 (2.02 to 3.29) |
| Middle East | 1.00 (reference) | 2.11 (1.78 to 2.51) | 2.34 (1.93 to 2.84) |
| North Africa | 1.00 (reference) | 1.36 (0.91 to 2.02) | 1.60 (1.03 to 2.50) |
| Subsaharan Africa | 1.00 (reference) | 1.88 (1.36 to 2.59) | 2.00 (1.41 to 2.84) |
| **Hospital contacts related to cardiopulmonary symptoms** |  |  |  |
| Denmark | 1.00 (reference) | 3.97 (3.91 to 4.04) | 4.10 (4.03 to 4.17) |
| Northern Europe | 1.00 (reference) | 3.68 (3.10 to 4.37) | 4.45 (3.68 to 5.37) |
| Western Europe | 1.00 (reference) | 3.40 (2.96 to 3.90) | 3.55 (3.06 to 4.12) |
| Eastern Europe | 1.00 (reference) | 4.00 (3.76 to 4.26) | 4.47 (4.17 to 4.80) |
| Asia | 1.00 (reference) | 3.51 (3.21 to 3.83) | 3.69 (3.35 to 4.07) |
| Middle East | 1.00 (reference) | 3.57 (3.31 to 3.83) | 4.31 (3.96 to 4.68) |
| North Africa | 1.00 (reference) | 3.99 (3.31 to 4.82) | 4.68 (3.81 to 5.74) |
| Subsaharan Africa | 1.00 (reference) | 2.68 (2.27 to 3.16) | 3.38 (2.81 to 4.07) |
| **Hospital contacts related to any long COVID symptoms** |  |  |  |
| Denmark | 1.00 (reference) | 4.14 (4.09 to 4.19) | 4.30 (4.25 to 4.36) |
| Northern Europe | 1.00 (reference) | 3.78 (3.33 to 4.28) | 4.36 (3.80 to 5.01) |
| Western Europe | 1.00 (reference) | 3.73 (3.37 to 4.14) | 3.92 (3.51 to 4.38) |
| Eastern Europe | 1.00 (reference) | 3.71 (3.54 to 3.89) | 4.15 (3.93 to 4.38) |
| Asia | 1.00 (reference) | 3.60 (3.36 to 3.85) | 3.71 (3.44 to 4.01) |
| Middle East | 1.00 (reference) | 3.24 (3.07 to 3.43) | 3.83 (3.59 to 4.08) |
| North Africa | 1.00 (reference) | 3.37 (2.93 to 3.86) | 4.07 (3.49 to 4.73) |
| Subsaharan Africa | 1.00 (reference) | 2.73 (2.41 to 3.09) | 3.29 (2.86 to 3.77) |

**Table S5.** Odds ratio of hospital contacts related to any long COVID symptoms 6 months after COVID-19 diagnosis compared with 6 months before COVID-19 diagnosis by largest countries of origin.

|  | **6 months before**<br>**COVID-19 diagnosis** | **6 months after COVID-19 diagnosis** |  |
| --- | --- | --- | --- |
|  | **OR (95% CI)** | **Unadjusted**<br>**OR (95% CI)** | **Adjusted**<br>**OR (95% CI)** |
| Denmark | 1.00 (reference) | 4.14 (4.09 to 4.19) | 4.30 (4.25 to 4.36) |
| Norway | 1.00 (reference) | 2.59 (2.08 to 3.21) | 2.69 (2.13 to 3.39) |
| Sweden | 1.00 (reference) | 5.22 (4.26 to 6.39) | 5.89 (4.70 to 7.37) |
| Afghanistan | 1.00 (reference) | 4.61 (3.95 to 5.37) | 4.67 (3.87 to 5.63) |
| Iraq | 1.00 (reference) | 3.04 (2.73 to 3.38) | 3.68 (3.25 to 4.17) |
| Iran | 1.00 (reference) | 4.52 (3.91 to 5.23) | 5.16 (4.40 to 6.04) |
| Somalia | 1.00 (reference) | 2.74 (2.31 to 3.26) | 3.24 (2.67 to 3.93) |
| Pakistan | 1.00 (reference) | 3.18 (2.84 to 3.57) | 3.38 (2.98 to 3.83) |
| Turkey | 1.00 (reference) | 3.50 (3.25 to 3.77) | 3.92 (3.61 to 4.25) |

**Table S6.** Odds ratios of hospital contacts related to any long COVID symptoms by largest countries of origin.

|  | 6 months before COVID-19 diagnosis |  |  | 0 to 4 weeks after COVID-19 diagnosis |  |  | >4 weeks to 6 months after COVID-19 diagnosis |  |  |
| --- | --- | --- | --- | --- | --- | --- | --- | --- | --- |
|  | n (%) | Unadjusted OR (95% CI) | Adjusted OR (95% CI) | n (%) | Unadjusted OR (95% CI) | Adjusted OR (95% CI) | n (%) | Unadjusted OR (95% CI) | Adjusted OR (95% CI) |
| Hospital contacts related to any long COVID symptoms* |  |  |  |  |  |  |  |  |  |
| Denmark | 25 375 (1.3) | 1.00 (reference) | 1.00 (reference) | 6506 (0.3) | 1.00 (reference) | 1.00 (reference) | 17 516 (0.9) | 1.00 (reference) | 1.00 (reference) |
| Norway | 83 (1.1) | 0.91 (0.79 to 1.05) | 0.88 (0.75 to 1.03) | 14 (0.2) | 0.74 (0.54 to 1.01) | 0.75 (0.54 to 1.04) | 62 (0.8) | 0.99 (0.83 to 1.17) | 1.04 (0.86 to 1.25) |
| Sweden | 81 (1.1) | 0.87 (0.74 to 1.01) | 0.81 (0.68 to 0.97) | 23 (0.3) | 0.73 (0.53 to 1.01) | 0.76 (0.54 to 1.07) | 81 (1.1) | 1.57 (1.36 to 1.80) | 1.74 (1.50 to 2.02) |
| Afghanistan | 178 (1.9) | 1.25 (1.11 to 1.40) | 1.04 (0.91 to 1.19) | 52 (0.5) | 1.73 (1.44 to 2.09) | 1.60 (1.28 to 1.99) | 170 (1.8) | 1.91 (1.70 to 2.14) | 1.31 (1.13 to 1.51) |
| Iraq | 382 (2.4) | 1.51 (1.40 to 1.62) | 1.19 (1.09 to 1.30) | 118 (0.7) | 1.73 (1.51 to 1.98) | 1.50 (1.29 to 1.75) | 280 (1.7) | 1.71 (1.57 to 1.87) | 1.26 (1.14 to 1.39) |
| Iran | 210 (2.2) | 1.57 (1.41 to 1.74) | 1.42 (1.27 to 1.59) | 60 (0.6) | 2.37 (2.01 to 2.79) | 2.26 (1.89 to 2.70) | 133 (1.4) | 1.79 (1.58 to 2.01) | 1.59 (1.40 to 1.81) |
| Somalia | 151 (1.6) | 1.38 (1.22 to 1.55) | 1.10 (0.96 to 1.26) | 47 (0.5) | 1.61 (1.30 to 2.00) | 1.51 (1.20 to 1.90) | 121 (1.3) | 1.68 (1.46 to 1.92) | 1.24 (1.06 to 1.44) |
| Pakistan | 278 (2.2) | 1.07 (0.98 to 1.16) | 0.94 (0.86 to 1.04) | 95 (0.7) | 1.43 (1.24 to 1.65) | 1.25 (1.07 to 1.47) | 217 (1.7) | 1.64 (1.51 to 1.79) | 1.37 (1.25 to 1.50) |
| Turkey | 749 (2.1) | 1.29 (1.23 to 1.37) | 1.07 (1.01 to 1.14) | 284 (0.8) | 2.06 (1.89 to 2.24) | 1.93 (1.75 to 2.12) | 666 (1.8) | 1.73 (1.63 to 1.83) | 1.32 (1.24 to 1.42) |

